# Impact of Atrial Fibrillation with Rapid Ventricular Response in Acute Decompensated Heart Failure

**DOI:** 10.1101/2024.06.20.24309212

**Authors:** Basheer Halhal, Omer Bar, Suliman Mohammad, Doron Aronson

## Abstract

**Background:** Rapid atrial fibrillation (AF) is considered to be a trigger for heart failure (HF) decompensation. Few data are available on AF (particularly with a rapid ventricular response) as a trigger for HF decompensation and its effect on clinical outcomes.

**Methods:** We studied 11,446 patients with acute HF. Rapid AF at admission was defined as ventricular rate ≥110 beats per minute, present at the first ECG performed after hospital arrival. The primary endpoint was in-hospital mortality.

**Results:** Rapid AF (defined as ventricular rate ≥ 110 beats per minute) at admission was present in 609 patients (5.3%). In-hospital mortality occurred in 83 (13.6%) and 951 (8.8%) patients with and without rapid AF, respectively. In a multivariable Cox regression model, the HR for in-hospital mortality was 1.50 (95% 1.16 to 1.93, *P*<0.0001). With further adjustment for heart rate, the effect of rapid AF on in-hospital mortality was no longer significant (HR 1.11; 95% CI 0.83–1.50, *P*=0.48).

From the original cohort, 419 participants with rapid AF in admission were matched on their propensity score to 419 patients with sinus tachycardia. In-hospital mortality occurred in 62 (14.8%) and 61 (14.6%) patients with and without rapid AF, respectively. Compared with the sinus tachycardia group, the HR for the in-hospital mortality in patients with rapid AF was 0.98 (95% CI 0.68 to 1.42; *P*=0.93).

**Conclusion:** Rapid AF in ADHF patients is associated with increased mortality risk that is mediated predominantly by rapid ventricular rate. The magnitude of the AF effect is similar to that of sinus tachycardia, indicating that the underlying mechanism for the adverse outcome is not directly related to AF.

## Introduction

Atrial fibrillation (AF) and heart failure (HF) often coexist, facilitating the occurrence and worsen the prognosis of each other.^1^ Epidemiological studies have demonstrated that AF often precedes the development of clinical HF,^2^ suggesting that AF can promote HF. However, whether AF is an independent predictor of adverse outcomes in HF or simply a marker of more advanced disease in sicker patients remains controversial.^3^

Observational data indicate that up to 40% of patients with ADHF who are admitted to the hospital have either previous AF, or new-onset AF at presentation.^4, 5^ Inappropriately high ventricular rates and irregularity of the cardiac rhythm can lead to symptoms or hemodynamic impairment, particularly in patients with elevated filling pressures. Therefore, AF is often considered to be a trigger for HF decompensation particularly if the ventricular response is not adequately controlled. However, although AF can precipitate HF decompensation in a previously stable patient, it is also possible that worsening HF has triggered an acute episode of AF or that previous AF with adequate rate control presents emergently with rapid ventricular rates due to HF decompensation.^6^

Few data are available on AF (particularly AF with rapid ventricular response) as a trigger for HF decompensation and its effect on clinical outcomes. Furthermore, prior studies have reported conflicting findings as to whether AF is an independent predictor of adverse outcomes in the setting of ADHF.^7–10^ Therefore, we studied the effect of AF with a rapid ventricular response on the outcome of patients with ADHF. Specifically, we tested whether the association between AF with a rapid ventricular response at admission and clinical outcome or detrimental hemodynamic changes in ADHF is mediated by the rapid heart rate or AF per se.

## Methods

We used a database of all patients admitted to our tertiary medical center with the primary diagnosis of AHF between January 2005 and Dec 2016. Eligible patients were those hospitalized with new-onset or worsening of preexisting heart failure as the primary cause of admission, using the European Society of Cardiology criteria.^11^ Patients were excluded if AHF was not the principal diagnosis during the admission. AF with a rapid ventricular response was defined as ventricular rate ≥ 110 beats per minute,^12^ present at the first ECG performed after hospital arrival.

The requirement for written informed consent was waived by the Rambam Hospital Institutional Review Board that gave its approval for this study.

### Endpoints

The primary endpoint was in-hospital mortality occurring up to 30 days from admission. The secondary endpoint was hemodynamic deterioration with need for inotrope or vasopressor therapy (dopamine, dobutamine, epinephrine, milrinone, norepinephrine, neosynephrin or vasopressin) within 48h from admission.

### Statistical analysis

Continuous variables are presented as mean = SD or medians (with interquartile ranges), and categorical variables as numbers and percentages. Baseline characteristics of the unmatched groups were compared using unpaired *t*-test for continuous variables and by the χ^2^ statistic for noncontinuous variables. After propensity score (PS) matching, the baseline covariates were compared between the two matched groups using a paired *t*-test or Wilcoxon signed rank test for continuous variables and the McNemar test or marginal homogeneity test for categorical variables.

The cumulative probability of the primary endpoint was assessed by the Kaplan-Meier method with significance testing by the log-rank statistic. Univariate and multivariable Cox proportional hazards regression models were used to analyze the relationship between rapid AF at admission and in-hospital mortality. All variables presented in Table 1 were considered as potential confounders. Variables perceived as clinically important based on prior knowledge and those with *P*<0.1 in univariable analysis were included in the multivariable Cox model.

**Table 1:** Baseline Clinical Characteristics.

| **Characteristics** | **Whole Study Population** |  |  | **Matched Groups** |  |  |
| --- | --- | --- | --- | --- | --- | --- |
|  | **No Rapid AF**<br>**(n = 10,837)** | **Rapid AF**<br>**(n = 609)** | **P value** | **Sinus**<br>**tachycardia**<br>**(n = 419)** | **Rapid AF**<br>**(n = 419)** | **P value** |
| Age (years) | 76 ± 12 | 77 ± 11 | 0.003 | 77 ± 11 | 77 ± 11 | 0.95 |
| Male | 5,351 (49) | 206 (34) | <0.0001 | 157 (37) | 144 (34) | 0.33 |
| Hypertension | 7,715 (71) | 451 (74) | 0.13 | 301 (72) | 309 (74) | 0.54 |
| Diabetes | 5,811 (54) | 282 (46) | <0.0001 | 197 (47) | 197 (47) | 1.00 |
| Chronic lung disease | 1,547 (14) | 76 (12) | 0.33 | 60 (14) | 54 (13) | 0.55 |
| Elevated troponin | 3,280 (30) | 196 (32) | 0.32 | 173 (41) | 164 (39) | 0.53 |
| Systolic blood pressure (mm Hg) | 146 ± 34 | 142 ± 28 | 0.01 | 142 ± 30 | 144 ± 29 | 0.39 |
| Diastolic blood pressure (mm Hg) | 76 ± 18 | 86 ± 18 | <0.0001 | 85 ± 19 | 86 ± 17 | 0.20 |
| Pulse pressure (mm Hg) | 69 ± 28 | 56 ± 20 | <0.0001 | 57 ± 20 | 57 ± 19 | 0.79 |
| Heart rate (beats/min) | 82 ± 18 | 125 ± 13 | <0.0001 | 123 ± 13 | 123 ± 13 | 0.56 |
| Estimated GFR (ml·min<sup>-1</sup>/1.73 m<sup>2</sup>) | 46 ± 24 | 48 ± 21 | 0.13 | 50 ± 21 | 48 ± 21 | 0.62 |
| Serum BUN (mg/dl) | 36 ± 22 | 32 ± 19 | 0.0001 | 33 ± 20 | 32 ± 20 | 0.53 |
| Glucose (mg/dL) | 175 ± 87 | 177 ± 91 | 0.58 | 187 ± 96 | 183 ± 96 | 0.54 |
| Albumin (g/dL) | 3.2 ± 0.5 | 3.1 ± 0.5 | 0.02 | 3.2 ± 0.6 | 3.2 ± 0.5 | 0.50 |
| Serum sodium (mmol/L) | 137 ± 5 | 137 ± 5 | 0.28 | 137 ± 5 | 137 ± 5 | 0.98 |
| Serum potassium (mmol/L) | 4.3 ± 0.7 | 4.2 ± 0.6 | 0.01 | 4.3 ± 0.7 | 4.3 ± 0.6 | 0.29 |
| Serum chloride (mmol/L) | 102 ± 5 | 102 ± 4 | 0.54 | 103 ± 5 | 102 ± 5 | 0.88 |
| Baseline hemoglobin (g/dL) | 11.4 ± 2.0 | 11.8 ± 1.8 | <0.0001 | 11.9 ± 1.9 | 11.8 ± 1.9 | 0.61 |
| Baseline hematocrit (%) | 35 ± 6 | 36 ± 5 | <0.0001 | 36 ± 6 | 36 ± 6 | 0.81 |
| White blood cell count | 10.4 ± 5.9 | 12.7 ± 11.0 | <0.0001 | 11.4 ± 6.9 | 11.1 ± 8.4 | 0.52 |
| BNP (pg/mL) | 836 [412–<br>1550] | 862 [512–<br>1374] | 0.78 | 828 [416–<br>1565] | 828 [467–<br>1361] | 0.59 |
| Fever (Temperature ≥38°C) | 245 (2) | 31 (5) | <0.0001 | 23 (5) | 18 (4) | 0.44 |
| **Left ventricular ejection**<br>**fraction\*** |  |  | 0.70 |  |  | 0.37 |
| Preserved ($\geq 50\%$) | 2,362 (47) | 118 (45) | | 71 (33) | 88 (48) | |
| Mid-range (41-49%) | 656 (13) | 31 (12) |  | 23 (11) | 19 (10) |  |
| Reduced ($\leq 40\%$) | 2,045 (40) | 112 (43) | | 121 (56) | 76 (42) | |
| **Medical therapy in hospital** |  |  |  |  |  |  |
| Beta blockers | 8,185 (76) | 468 (77) | 0.46 | 313 (75) | 321 (77) | 0.52 |
| ACEi/A-II blockers | 7,262 (67) | 387 (63) | 0.08 | 263 (63) | 263 (63) | 1.00 |
| Loop diuretics | 7,446 (69) | 412 (68) | 0.58 | 275 (66) | 283 (68) | 0.55 |
| MRA | 1,404 (13) | 57 (9) | 0.01 | 46 (11) | 39 (9) | 0.43 |

Data are number (%) or mean ± SD
\* Available in 47% of patients

Because the clinical characteristics of patients with rapid AF differed from those without, results of the multivariable analysis were verified using propensity score matching. Propensity score estimates representing the probability of rapid AF at admission were generated using a non-parsimonious multiple logistic regression model derived from baseline clinical and laboratory covariates (Table 1). Following propensity score generation, patients were matched by using 1:1 nearest neighbor (Greedy type) matching without replacement and a caliper width of a standard deviation of the propensity score logit. Matching was performed without replacement, and nonmatched results were discarded.

We assessed the success of the matches by examining standardized differences (measured in percentage points) in the observed confounders between the matched groups. Small (<10%) standardized differences support the assumption of balance between groups based on observed confounders.^13^ After matching, the Cox proportional hazards models were stratified by the pair to account for dependence among matched subjects.^13^ Statistical analyses were performed using Stata version 18.0 (Stata Corp., College Station, TX, USA).

## Results

Between January 2006 and December 2016, a total of 11,446 patients were enrolled in the study. Of these, 609 patients (5.3%) presented with rapid AF. The baseline clinical and characteristics of the study population according to the presence of rapid AF at admission are shown in Table 1 (right panel). Patients with rapid AF were older, more likely to be females, present with lower systolic blood pressure and pulse pressure, fever and higher white blood cell count. There were no differences in baseline medical therapies between the groups. Length of hospital stay was longer in the rapid AF group (median 6 days [IQR 4 to 11] vs 5 days [IQR 3 to 9], *P*<0.0001).

When cubic spline regression was used to explore the association between heart rate at admission (irrespective of baseline rhythm) and in-hospital mortality, we observed a near linear increase in the risk for mortality with increasing heart rate (Figure 1) irrespective of baseline rhythm.

**Figure 1:**
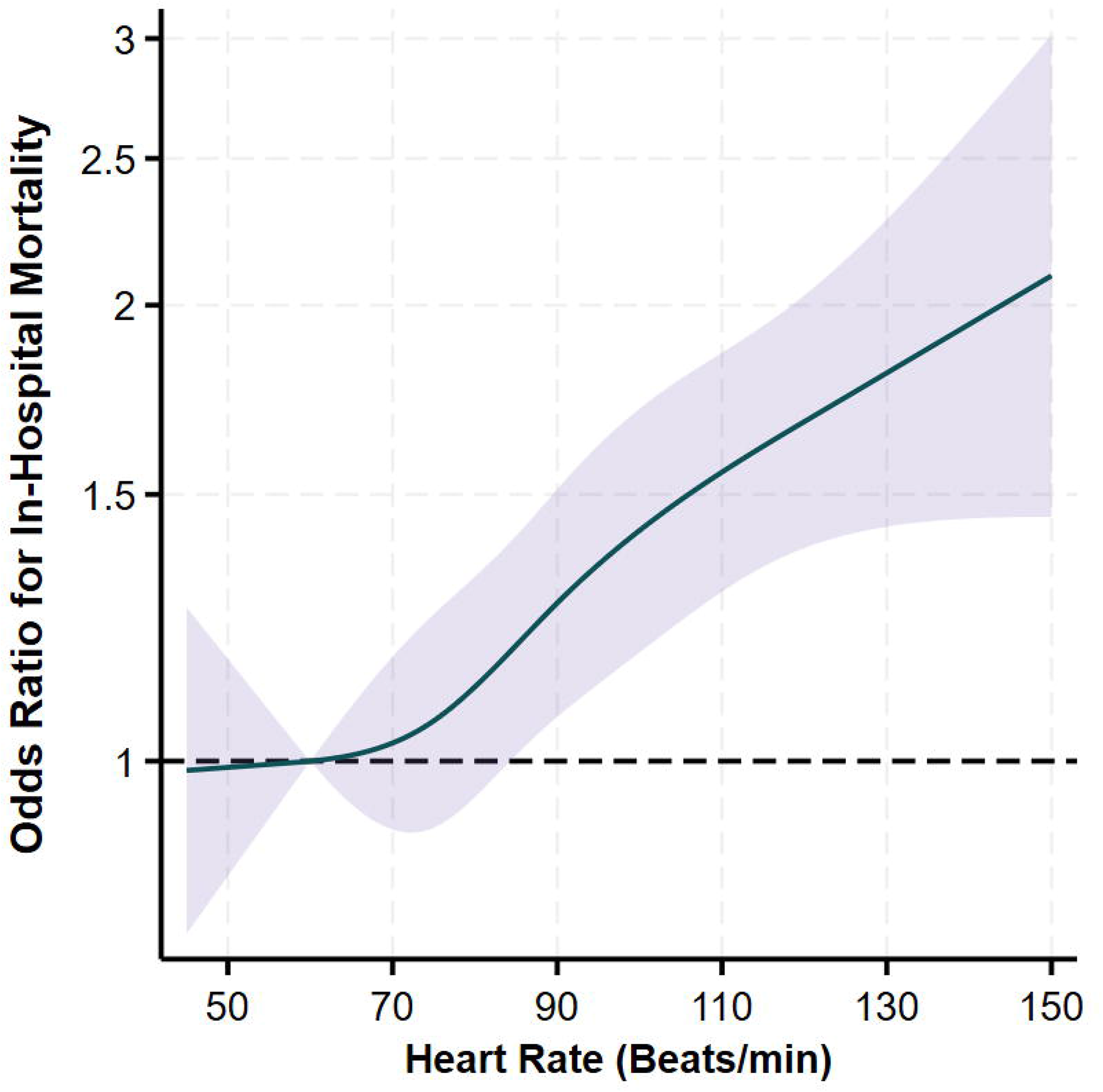
**A.** Predicted probability of in-hospital mortality according to heart rate at admission. **B.** Spline function graph of the adjusted relation between heart rate and in-hospital mortality.

### Changes in heart rate during hospitalization

Figure 2 depicts the changes in heart rate during the first 7 days of hospitalization in patients presenting with rapid AF, patients presenting with sinus tachycardia and patients presenting without tachycardia. In patients presenting with rapid AF or sinus tachycardia, the heart rate decreased predominantly during the first 24 hours, followed by a slower decrease at later timepoints (Figure 2). The majority of patients with rapid AF were treated medically and only 25 (4.1%) underwent cardioversion.

**Figure 2:**
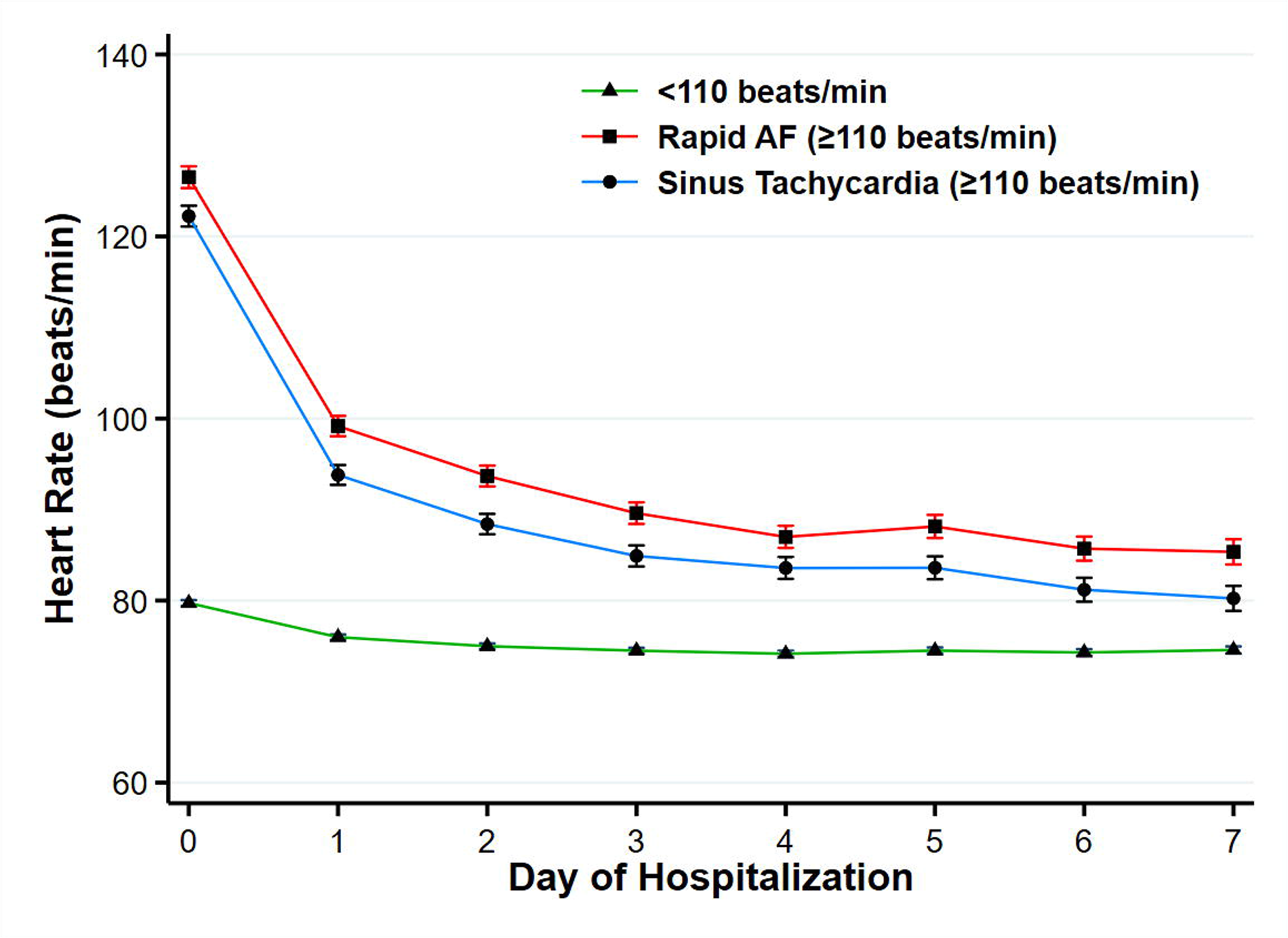
Changes in heart rate during the first 7 days of hospitalization in patients classified according to heart rate at admission (rapid atrial fibrillation, sinus tachycardia and without rapid heart rate).

In a linear mixed model, there was no significant change in the heart rate slope between patients presenting with rapid AF and sinus tachycardia (*P*=0.24). However, compared with patients without tachycardia at baseline, the heart rate of patients presenting with rapid AF or sinus tachycardia remained significantly higher throughout the hospitalization (*P*<0.0001 for both comparisons).

### Rapid AF and in-hospital mortality

In-hospital mortality occurred in 83 (13.6%) and 951 (8.8%) patients with and without rapid AF, respectively (**Figure 3**A). In an unadjusted Cox regression model, the HR for in-hospital mortality was 1.59 (95% 1.29 to 1.99, *P*<0.0001). Adjustments for multiple risk factors in a multivariable Cox proportional hazards regression model had a small effect on the hazard of in-hospital mortality (Adjusted HR 1.50; 95% CI 1.16– 1.93, *P*=0.002). However, with further adjustment for heart rate, the effect of rapid AF on in-hospital mortality was no longer significant (HR 1.11; 95% CI 0.83–1.50, *P*=0.48).

**Figure 3:**
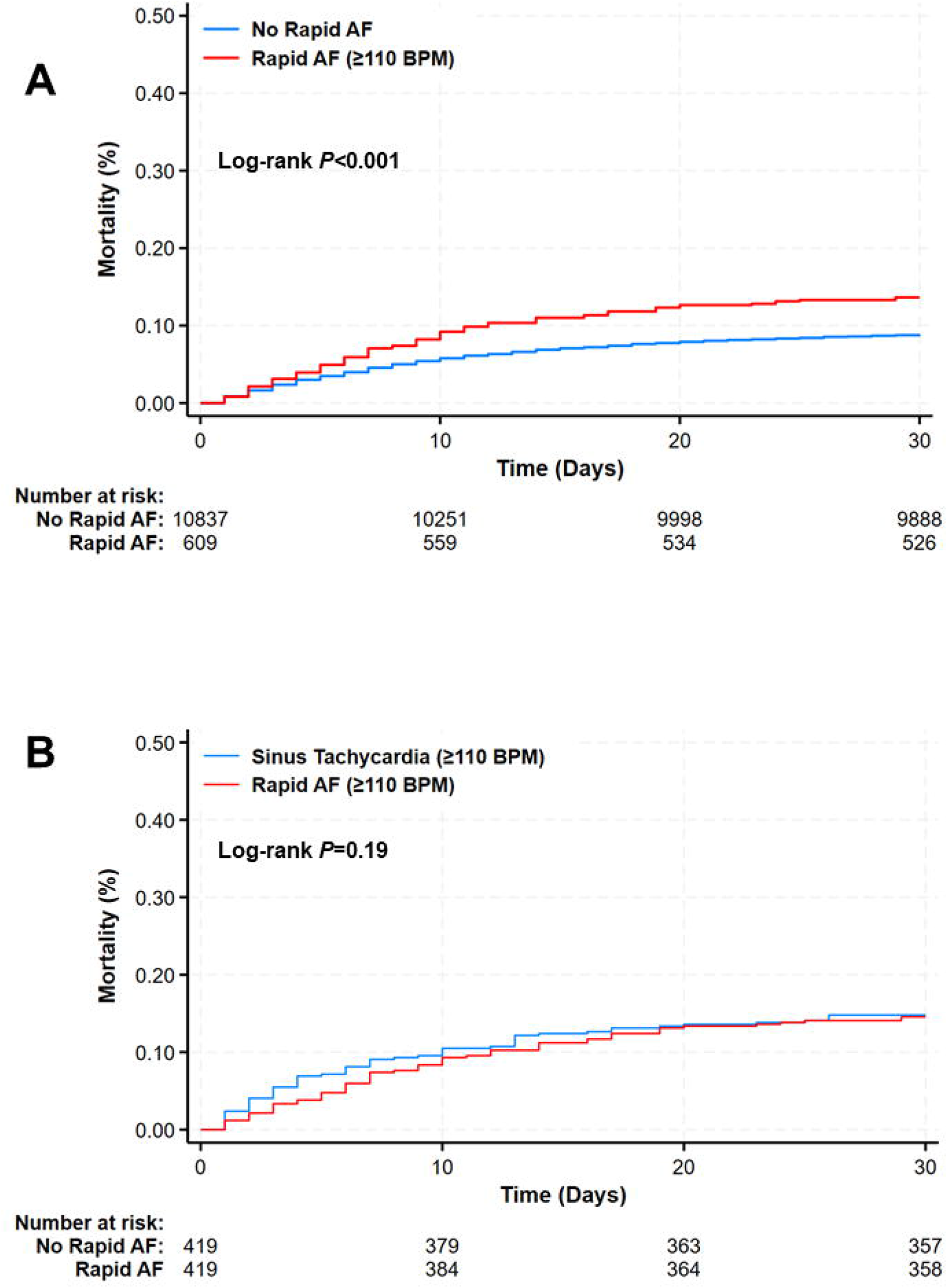
In hospital mortality up to 30 days according to rapid atrial fibrillation at admission. **(A)** Unadjusted in the whole population and **(B)** After propensity score matching of patients with rapid atrial fibrillation and sinus tachycardia.

Compared with patients without increased heart rate at admission (n=10,100), both patients with sinus tachycardia (heart rate ≥ 110 BPM, n=737) and rapid AF (AF heart rate ≥110 BPM, n=609) had an increased risk of in-hospital mortality (HR 1.44, 95% CI 1.15–1.81 and 1.51, 95% CI 1.19–1.92, respectively). However, in hospital mortality was similar among patients with sinus tachycardia and rapid AF (HR 1.04, 95% CI 0.76–1.43; Figure 4).

**Figure 4:**
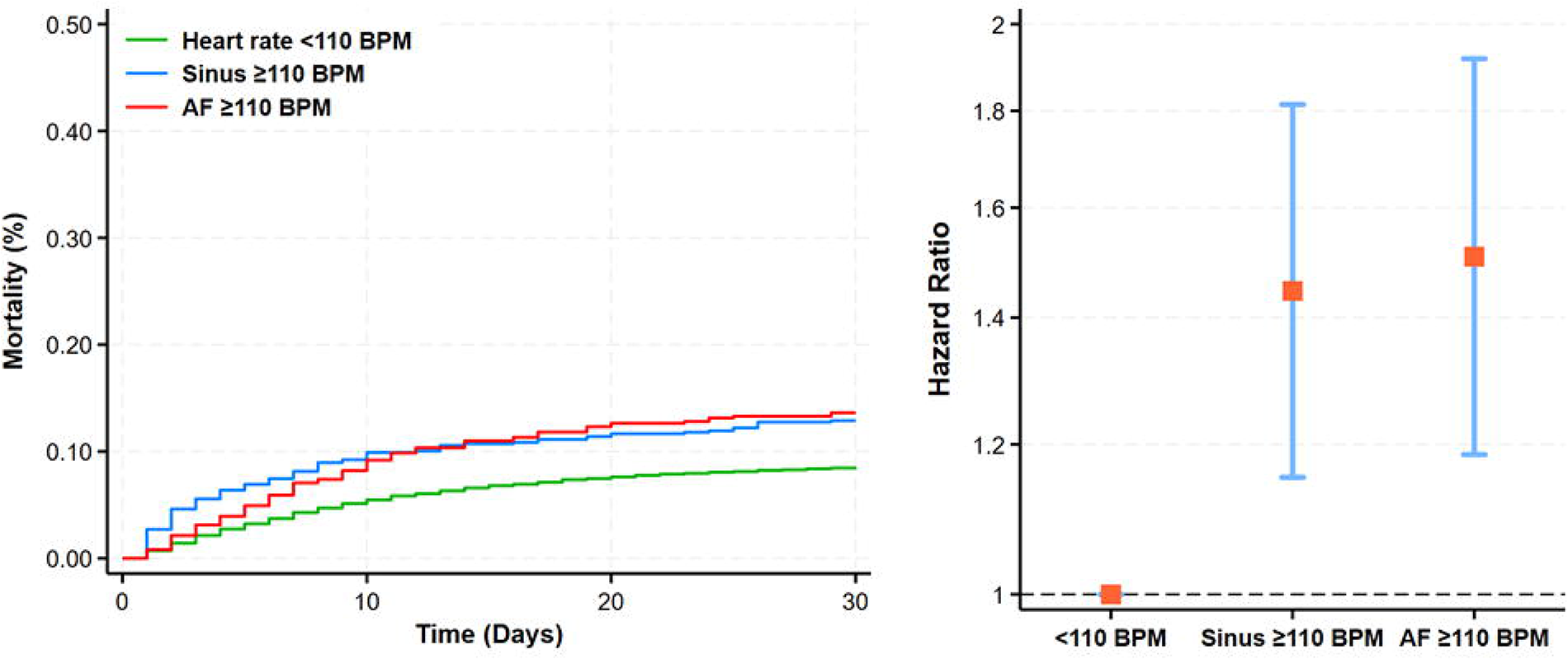
**(A)** Kaplan–Meier curves showing cumulative incidence of atrial fibrillation according to heart rate status (*P*<0.0001, log-rank test). (B) Adjusted hazard ratios and 95% confidence intervals for each category.

### Propensity score matching

From the original cohort, 419 participants with rapid AF in admission were matched on their propensity score to 419 patients with sinus rhythm (sinus tachycardia). After propensity score matching, the mean standardized difference in covariates between the two groups decreased from 21.4% before matching to 3.3% after matching.

Patients were well balanced with respect to the individual variables included in the propensity model, with absolute standard differences between <10% for all variables (Figure 5). In the matched cohort, there were no significant differences between the groups for all clinical characteristics (Table 1, Right panel).

**Figure 5:**
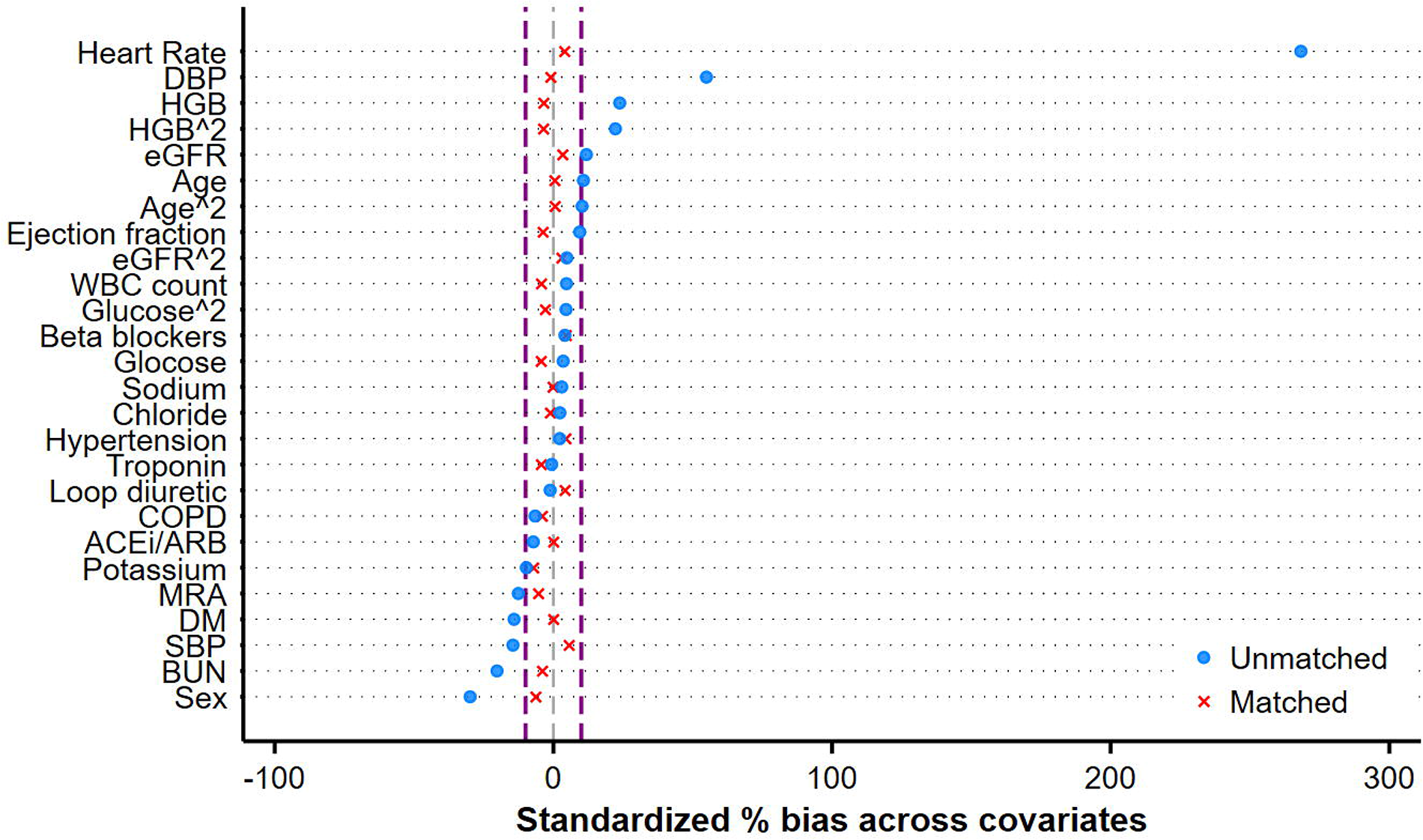
Covariable balance before (red circles) and after (green exes) matching. The standardized difference after propensity matching (blue lines) are all well within 10%.

Following propensity score matching, there were 61 in-hospital mortality events (14.6%) in patients with rapid AF and 62 events (14.8%) in patients without rapid AF (**Figure 3** B). Compared with the control (sinus tachycardia) group, the HR for the in-hospital mortality in patients with rapid AF was 0.98 (95% CI 0.68 to 1.42; *P*=0.93).

#### Rapid AF and hemodynamic instability

Hemodynamic instability in the first 48h of hospitalization occurred in 35 (5.8%) and 339 (3.1%) patients with and without rapid AF, respectively (*P*<0.0001). In an unadjusted logistic regression model, rapid AF was associated with an unadjusted odds ratio (OR) of 1.88 (95% 1.32 to 2.70, *P*=0.001) for hemodynamic instability. Adjustments for multiple risk factors in a multivariable logistic regression modestly mitigated the odds of hemodynamic instability (Adjusted OR 1.67; 95% CI 1.10–2.53, *P*=0.017). However, after adjusting for heart rate, the association of rapid AF with hemodynamic instability was no longer significant (OR 1.09; 95% CI 0.66–1.80, *P*=0.73).

## Discussion

The goal of the present study was to examine the clinical consequences of AF with a rapid ventricular response in patients with ADHF. Rapid AF at admission was associated with an increased risk for in-hospital mortality and hemodynamic deterioration within 48h. However, this association was no longer significant after adjustment for heart rate. Furthermore, propensity score matching of patients with rapid AF with patients with sinus tachycardia at admission demonstrated no difference in mortality risk. Overall, these findings suggest that the main deleterious effect of AF with a rapid ventricular response is mediated via the high heart rate, with the magnitude of risk being similar to that of sinus tachycardia.

A major concern with rapid ventricular response in AF is the worsening of HF symptoms.^2^ However, surprisingly, there is little evidence that rapid AF adversely affects the outcome of patients with AHF.^7–10^ In the intercontinental GREAT registry, AHF precipitated by AF showed lower 90-day risk of death compared with AHF without identified precipitants (HR 0.56, 95% CI 0.42–0.75, P < 0.001).^8^ These observations are in line with those of the Spanish PAPRICA-2 (PApel pronostico de los PRecipitantes de un episodio de Insuficiencia Cardiaca Aguda) study, which showed relatively favorable short-term outcome when the AHF was judged to be precipitated by AF.^9^ In the Korean Acute Heart Failure registry, patients whose acute decompensation was classified as tachycardia-mediated had better in-hospital outcome, and similar post-discharge outcomes compared with those with sinus rhythm or those with AF whose decompensation was not considered to be tachycardia-mediated.^10^ However, in these studies, there was no clear definition for classifying the decompensation event as AF or tachycardia-mediated.

By contrast, in the EuroHeart Failure Survey, new-onset AF (but not previous AF) was an independent predictor of in-hospital mortality (odds ratio 1.53, 95% CI 1.1–2.0).^5^ The discrepancy between some of these studies and the current report is probably related to the fact that the ventricular rate at admission was not considered and lack of adjustments for heart rate.

### Worsening of HF and AF

Several mechanisms may explain why AF can exacerbate HF. First, AF with persistent rapid ventricular response can lead to ventricular impairment with progressive dilatation and increase in LV wall stress and end-diastolic pressure and volume and reduced myocardial perfusion in association with cellular effects such as mitochondrial disruption, abnormal calcium handling and cardiac inflammation.^14–16^ Second, the loss of atrial booster pump function may predispose to HF by causing a fall in cardiac output.^17^ Third, several studies suggest that heart rate irregularity per se (irregularity-induced cardiomyopathy) with calcium mishandling can lead to left ventricular dysfunction despite appropriate rate control.^15, 18^ The current study therefore suggests that increased heart rate is the predominant mechanism that can contribute to HF decompensation and increase short term mortality.

In patients presenting with AHF and AF with a rapid ventricular response, either the AF episode itself triggered decompensation in a previously stable patient or worsening HF has precipitated an acute episode of AF.^6^ Although it is difficult to establish a causal link between the AF and the decompensation and the clinical impression that AF triggers AHF is subjective, it is commonly accepted and inferred in the presence of a rapid ventricular response. Hance, acute rate control and potentially acute rhythm management is often considered. Emergency electrical cardioversion is recommended in AF patients with acute or worsening hemodynamic instability (class IB^1^ or IC^19^), including acute pulmonary oedema, symptomatic hypotension, or cardiogenic shock.^1^ Although cardioversion may transiently restore sinus rhythm, the expected recurrence rates in the still-decompensated patient is very high.^6, 20^ In more stable patients, congestion relief may reduce sympathetic drive and ventricular rate and increase the probability of spontaneous return to sinus rhythm.^19^

Conversely, HF may predispose to AF via several mechanisms. Experimental and clinical observations suggest that increasing atrial pressure and/or acute atrial dilatation or increased atrial stretch induced by increased atrial pressure shortens atrial refractory period and greatly increases the vulnerability to AF.^21^ In animal models, acute atrial dilatation induced by increased atrial pressure produced shortening of atrial effective refractory period with increased vulnerability to AF,^21, 22^ through stimulation of atrial stretch-activated ion channels.^21, 23, 24^ Chronically, HF causes atrial fibrosis and regional conduction abnormalities, providing a substrate for AF initiation and maitence.^24^ Furthermore, the sympathetic activation seen in AHF may contribute to electrophysiologic changes, such as a shortened atrial refractory period, that promote AF with a rapid ventricular response.^25^ Therefore, it is also possible that the cause of new-onset AF is also the reason for the increase in mortality, making rapid AF a marker but not necessarily causally inked to mortality.

In the present study, increased heart rate on admission for AHF was independently associated with in-hospital mortality, as previously reported in chronic HF.^26, 27^ A lower resting heart rate is typically a marker of intrinsically reduced sympathetic tone, which consequently indicates lower HF severity and risk. The observation that sinus tachycardia carries a risk similar to that of rapid AF suggests that increased sympathetic tone, indicative of more severe disease or other triggers for decompensation, is an underlying mechanism leading to either sinus tachycardia or rapid AF through accelerated AV conduction. These underlying processes likely impact the outcome more significantly than the secondary phenomena of rapid AF.

### Study limitations

It is important to consider several limitations pertinent to the methods of this study. This was a single-center post-hoc analysis and thus, the results must be regarded as hypothesis generating and exploratory and require validation in other studies. The duration of the rapid AF prior to hospital admission could not be ascertained. This may be an important factor of clinical outcome that could have affected our risk estimates. We did not consider rapid AF episodes that developed after admission. The impact of rapid AF on left ventricular systolic function was not assessed.

### Conclusion

Rapid AF in ADHF patients is associated with increased mortality and hemodynamic deterioration that is mediated predominantly by the rapid ventricular response. The magnitude of the AF effect on mortality is comparable to that of sinus tachycardia, suggesting that the adverse effects are not directly caused by AF itself.

## Data Availability

All data produced in the present study are available upon reasonable request to the authors

